# Emergence of a novel SARS-CoV-2 strain in Southern California, USA

**DOI:** 10.1101/2021.01.18.21249786

**Authors:** Wenjuan Zhang, Brian D Davis, Stephanie S Chen, Jorge M Sincuir Martinez, Jasmine T Plummer, Eric Vail

## Abstract

Since October 2020, novel strains of SARS-CoV-2 including B.1.1.7, have been identified to be of global significance from an infection and surveillance perspective. While this strain (B.1.1.7) may play an important role in increased COVID rates in the UK, there are still no reported strains to account for the spike of cases in Los Angeles (LA) and California as a whole, which currently has some of the highest absolute and per-capita COVID transmission rates in the country. From the early days of the pandemic when LA only had a single viral genome uploaded onto GISAID we have been at the forefront of generating and analyzing the SARS-CoV-2 sequencing data from the LA region. We report a novel strain emerging in Southern California. Most current cases in the catchment population in LA fall into two distinct subclades: 1) 20G (24% of total) is the predominant subclade currently in the United States 2) a relatively novel strain in clade 20C, CAL.20C strain (∼36% of total) is defined by five concurrent mutations. After an analysis of all of the publicly available data and a comparison to our recent sequences, we see a dramatic growth in the relative percentage of the CAL.20C strain beginning in November of 2020. The predominance of this strain coincides with the increased positivity rate seen in this region. Unlike 20G, this novel strain CAL.20C is defined by multiple mutations in the S protein, a characteristic it shares with both the UK and South African strains, both of which are of significant clinical and scientific interest

## Introduction

The first systematic genetic analysis of SARS-CoV-2 from Southern California showed most isolates originated from clade 20C that likely emerged from New York via Europe early in the COVID-19 pandemic(1). Since October 2020, novel strains of SARS-CoV-2 including B.1.1.7(UK) and B.1.351(South Africa) have become globally important because of their local dominance in COVID-19 positive cases(2). While the B.1.1.7 strain may play an important role in increased COVID rates in the UK and Europe, there are still no reports to account for the current spike of cases in Los Angeles and California as a whole that began in early November 2020. We report the existence of a novel strain CAL.20C that is currently increasing in numbers in Southern California.

## Methods

Regulatory review was completed by the CSMC Office of Research Compliance and Quality Improvement (IRB # STUDY629). In total, 192 SARS-CoV-2 positive nasopharyngeal samples collected between 11/22/2020 and 12/28/2020 were processed for NGS and as per our protocol (1) with the following updates. Total RNA(100ng) was processed for cDNA synthesis using the Illumina RNA preparation enrichment tagmentation kit. Sequencing libraries were made using 200 ng of cDNA and a viral respiratory panel(RVOPV2). Samples were pooled and sequenced on an Illumina NovaSeq(2×100 bp). All sequencing reads were mapped to SARS-CoV-2 genome and data deposited to GISAID(3). This data was combined with all publicly available sequences(N = 4,337) from Southern California(Imperial, Kern, Los Angeles, Orange, Riverside, San Bernardino, San Diego, San Luis Obispo, Santa Barbara, and Ventura Counties) and phylogenetic analysis was performed as previously reported(1).

## Results

We detected a novel strain descended from cluster 20C and defined by five mutations (ORF1a: I4205V, ORF1b:D1183Y, S: S13I;W152C;L452R)(Figure 1). This strain, CAL.20C, was first observed in July 2020 in 1/1230 samples from LA county and not detected in Southern California again until October. Since then, this strain’s prevalence has increased absolutely and relatively in Southern California, where by December it accounted for 24% of all samples (Figure 2A) and 36.4% (66/181) of our local Los Angeles cohort. The emerging predominance of this strain temporally tracks to a time at or before the onset of the current spike in Southern California(Figure 2B). Unlike clade 20G, which is currently the largest reported clade in North America, this clade is defined by multiple mutations in the S protein (similar to B.1.1.7/B.1.351) and represents a separate subclade of 20C (Figure 3). Currently the CAL.20C strain is primarily found in Southern California, however we have detected multiple recent isolates in Northern California, New York and Washington DC. Additionally, our analysis reveals a small number of cases found outside of the US in Oceania in the past month.

**Figure 1.**
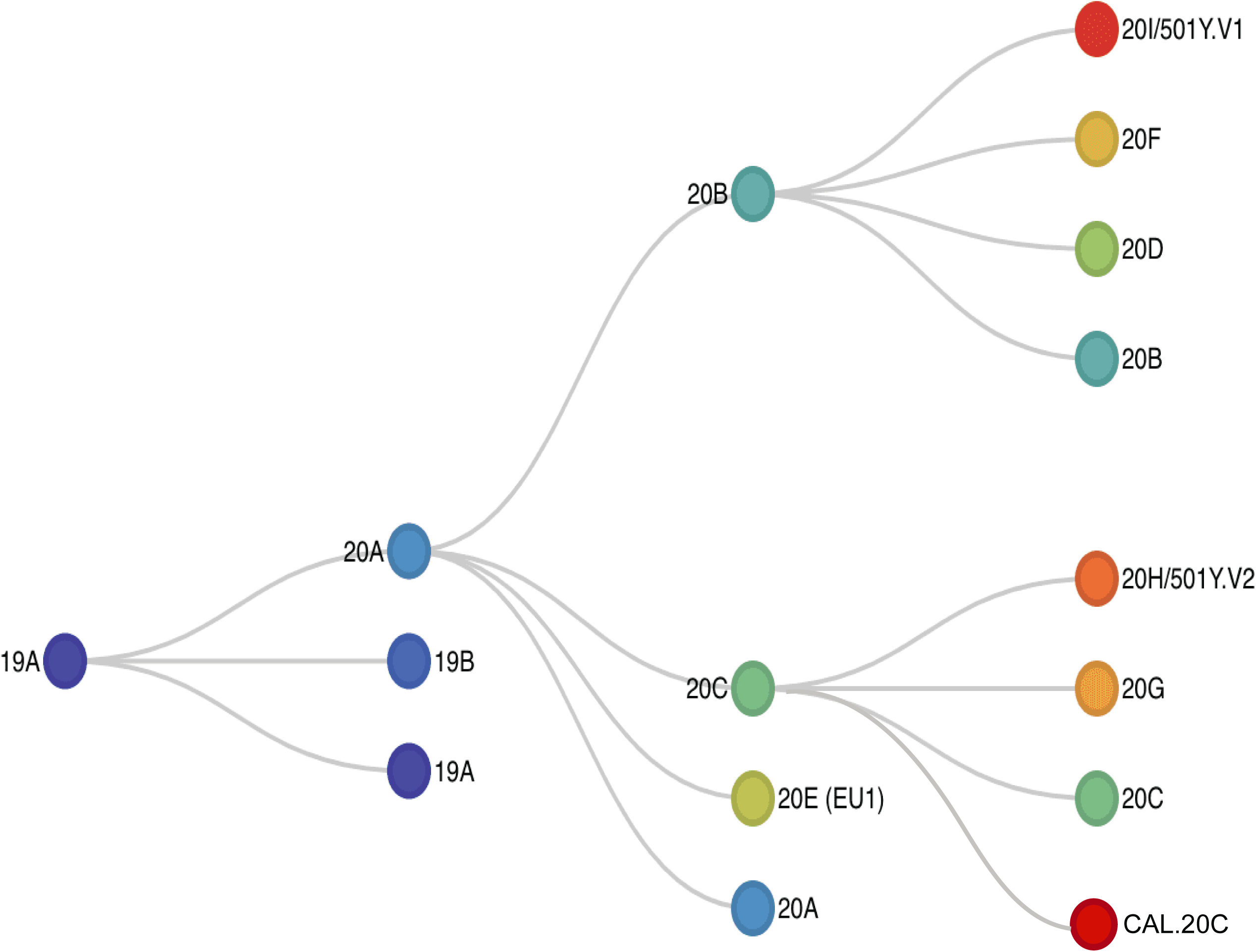
Phylogenetic classification of SARS-CoV-2 genomes. Phylogenetic analysis using 192 Los Angeles isolates and a global subsampling from May 2020 to January 2021 reveal a novel Southern California strain CAL.20C.

**Figure 2.**
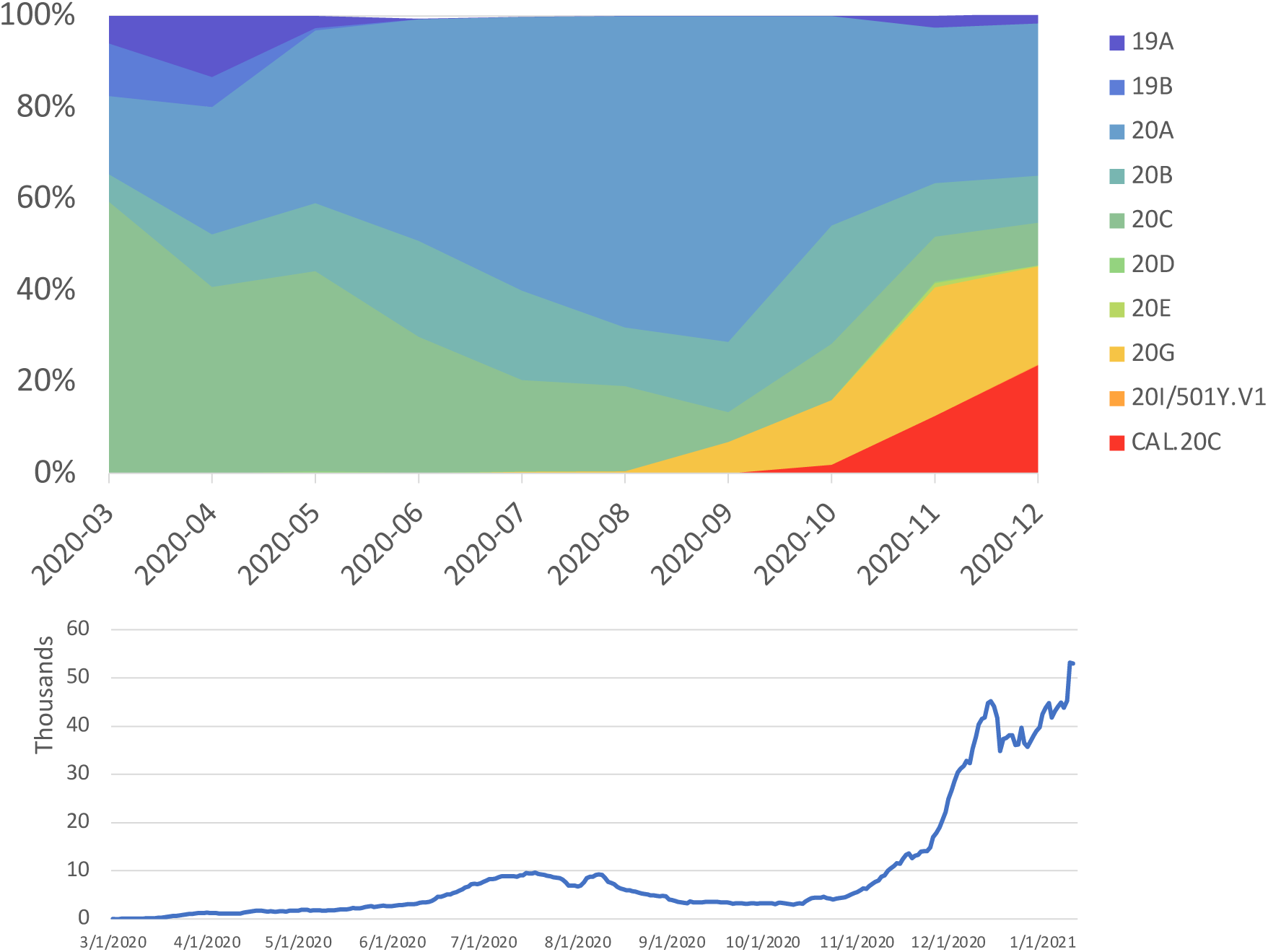
Timeline for the emergence of a novel Southern California strain, CAL.20C, amongst all SARS-CoV-2 strains observed. a) Diagrammatic representation of isolates from Southern California corresponding to collection time points spanning emergence of first isolates (3/2020) to present COVID positive cases (12/2020). b) 7 day average of confirmed COVID-19 cases in the Southern California region.

**Figure 3.**
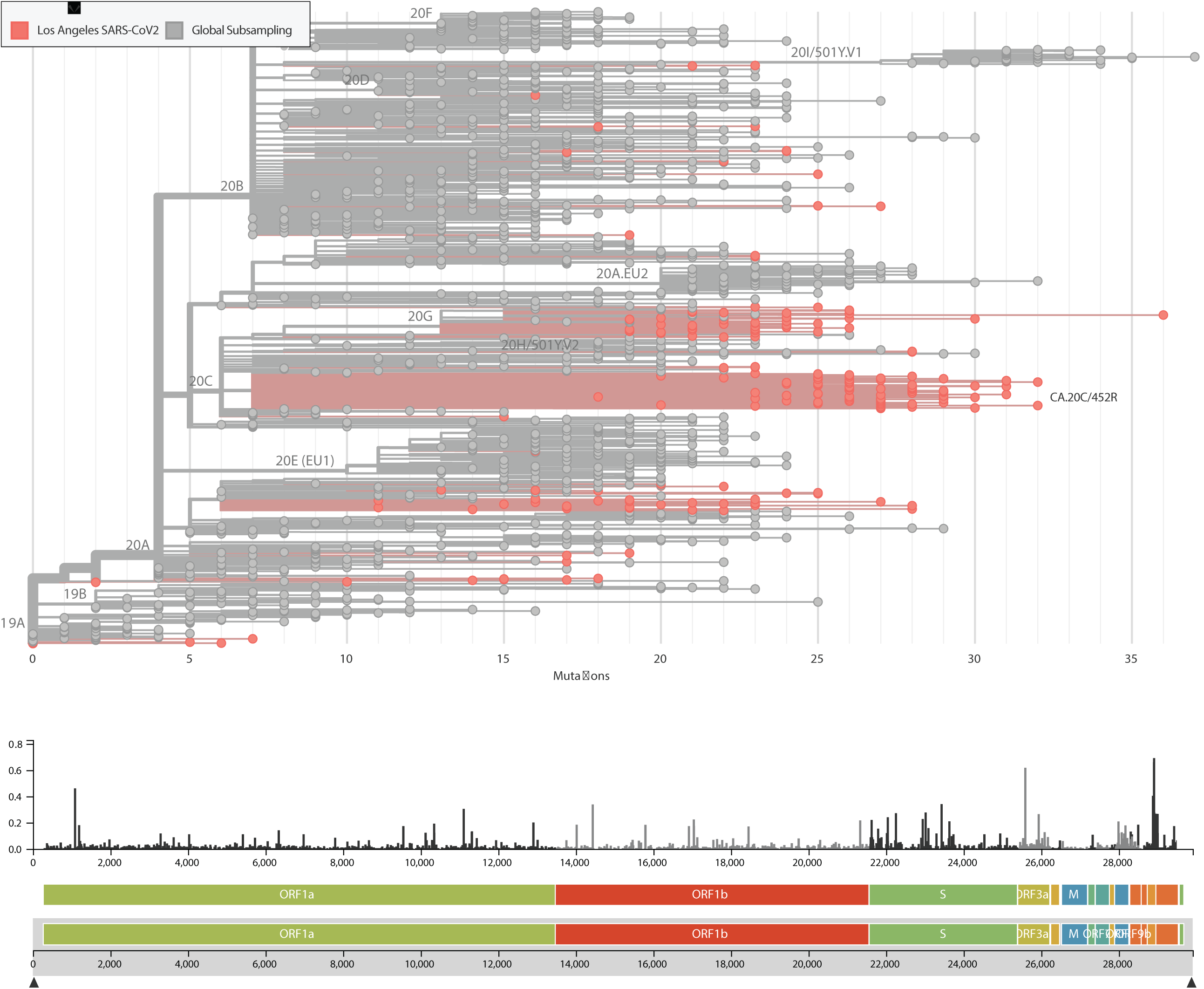
Phylogenetic tree of SARS-CoV-2 genomes.

## Discussion

These findings add to our current understanding of COVID-19 transmission within the US, specifically that the recent surge in COVID-19 positive cases in Southern California coincides with the emergence of a unique strain, CAL.20C. Given the independent emergence of geographically isolated, clinically relevant strains such as B.1.1.7 (UK) and B.1.351 (South Africa), the CAL.20C strain may be partially responsible for the magnitude of the surge in COVID-19 on the West Coast of the US. The S protein L452R mutation is within a known receptor binding domain that has been found to be markedly resistant to certain monoclonal antibodies to the spike protein(4). Mutations in this domain may be resistant to polyclonal sera as seen in convalescent patients or those post vaccination(5). The functional effect of this mutation in concert with other detected mutations in CAL.20C, both in terms of infectivity and antibody resistance is unknown at this time. The identification of this novel strain is important to frontline and global surveillance of this constantly evolving virus and has a direct impact on public health, especially vaccination efforts.

## Data Availability

All data has been deposited to GISAID

